# Evaluation Of The *Rims2* Locus As A Risk Locus For Parkinson’s Disease Dementia

**DOI:** 10.1101/2021.07.26.21261131

**Authors:** MX Tan Manuela, Real Raquel, A Lawton Michael, Bresner Catherine, Sofia Kanavou, Thomas Foltynie, W Wood Nicholas, A. Nalls Mike, M Williams Nigel, Ben-Shlomo Yoav, TM Hu Michele, G Grosset Donald, Hardy John, Shoai Maryam, R Morris Huw

## Abstract

In their recent letter entitled “Genome-wide survival study identifies a novel synaptic locus and polygenic score for cognitive progression in parkinson’s disease”, Liu and colleagues report that in a genome-wide analysis of progression to dementia in parkinson’s disease (PD) the *RIMS2* locus is a determinant of dementia in PD (1). In this study we have evaluated the nominated loci in a well-powered longitudinal clinical-genetic study of 2536 individuals in the tracking parkinson’s and oxford discovery cohorts. We have not identified any association between the *RIMS2* locus or other loci from the discovery phase and the development of Parkinson’s dementia. further work is needed to understand the biological determinants of this important aspect of parkinson’s and to guide the search for new treatments.

**Brief paragraph/Abstract:** In their recent letter entitled “Genome-wide survival study identifies a novel synaptic locus and polygenic score for cognitive progression in Parkinson’s disease”, Liu and colleagues report that in a genome-wide analysis of progression to dementia in Parkinson’s disease (PD) the *RIMS2* locus is a determinant of dementia in PD (1). In this study we have evaluated the nominated loci in a well-powered longitudinal clinical-genetic study of 2536 individuals in the Tracking Parkinson’s and Oxford Discovery cohorts. We have not identified any association between the *RIMS2* locus or other loci from the discovery phase and the development of Parkinson’s dementia. Further work is needed to understand the biological determinants of this important aspect of Parkinson’s and to guide the search for new treatments.

## Main text

This work is important since genetic determinants of dementia in PD may be important for understanding underlying disease biology, stratifying patient groups and developing new treatments. Liu and colleagues studied 4,872 patients from 15 cohorts, with varying definitions of dementia. They used 11.2 million imputed single nucleotide variants (SNVs), derived from the 1.8 million variants genotyped on the high density Illumina Infinium Multi-Ethnic Global Array. Imputed variants with a minor allele frequency of ≥ 0.1% and an R^2^ of ≥ 0.3 were included. Using a single pooled genome-wide Cox proportional hazards approach with individual studies as a covariate, they identified nine SNVs which passed genome-wide significance in the discovery phase (*P*-value < 5 × 10^−8^), with an intronic SNV within the *RIMS2* gene achieving nominal significance in the replication phase (p=0.004) to provide a combined hazard ratio (HR) of 4.74 and *P*-value of 2.78 × 10^−11^. The *RIMS2* SNV has a rare allele frequency of 1.3%, lies around 6 kb from exon 2 of *RIMS2* (NM_001282882.2, transcript variant 4) and has not been reported to be in linkage disequilibrium with a coding variant, nor to act as an expression or splicing QTL for transcripts in this region. The significant signal at the *RIMS2* locus relates to ∼ 90/3735 SNV variant carriers. Interestingly, *GBA* and *ApoE* did not achieve genome-wide significance for time-to-dementia in PD but were nominally significant in a candidate gene analysis of eight genes.

Two previous large genome-wide studies of the genetic determinants of dementia in PD have not identified the *RIMS2* locus as relevant to PD dementia (2,3), although one of these studies used a minor allele frequency filter of > 5% and may have missed the effect of rare alleles(2,3).

We have evaluated serial cognitive data in 2,536 PD patients from the Tracking Parkinson’s and Oxford Discovery cohorts (4,5), two large United Kingdom based studies of the genetic determinants of dementia in PD using almost identical study protocols. In both studies, dementia was defined as a Montreal Cognitive Assessment (MoCA) score of ≤ 21 or study withdrawal due to a diagnosis of dementia. Genotyping was carried out on the Illumina HumanCoreExome-12 v1.1 and Illumina Infinium HumanCoreExome-24 v1.1 array (Oxford Discovery) and Illumina HumanCoreExome array with custom content (Tracking Parkinson’s). Imputation was carried out using the Michigan Imputation server against the 1000 Genomes Project reference panel.

Twenty eight percent (710/2,536) of the combined UK cohorts met the endpoint of dementia. Cox proportional hazards analysis was carried out evaluating each of the nine SNVs nominated by Liu and colleagues, and an informative proxy SNP, for time from motor onset to dementia, adjusting for age at onset, sex, and the first ten principal components (Protocol: Protocols.io). Analyses were conducted in each cohort separately and then meta-analysed using random-effects meta-analysis. A conservative random effects meta-analysis approach was chosen because of differences in the baseline symptom severity. Power calculations were carried out for time-to-event analysis using the R package survSNP, using an alpha of 5 × 10^−8^ and the event time of median time to dementia of 4.0 years (4.2 years in Tracking Parkinson’s and 3.9 years in Oxford Discovery). With a HR ratio of 4.74 we have 99.7% power in the combined analysis of Tracking Parkinson’s and Oxford Discovery to detect an effect of the risk allele on the risk of developing dementia.

We did not identify a significant effect of any of the nominated discovery SNVs on the risk of developing dementia (Table), including the *RIMS2* locus, either in the discovery SNV or in high quality (INFO score> 0.3, r^2^ > 0.35) proxy SNVs. Two SNVs in *RP11-705O24*.*1* were nominally significant following a heavily reduced Bonferroni correction for multiple independent tests (rs118029233 and rs118004610) in the Oxford Discovery cohort evaluating 18 SNPs. However, this was not replicated in the Liu study, in the Tracking Parkinson’s cohort, or in our meta-analysis, meaning these variants are unlikely to be a risk factor for PD dementia.

**Table.**
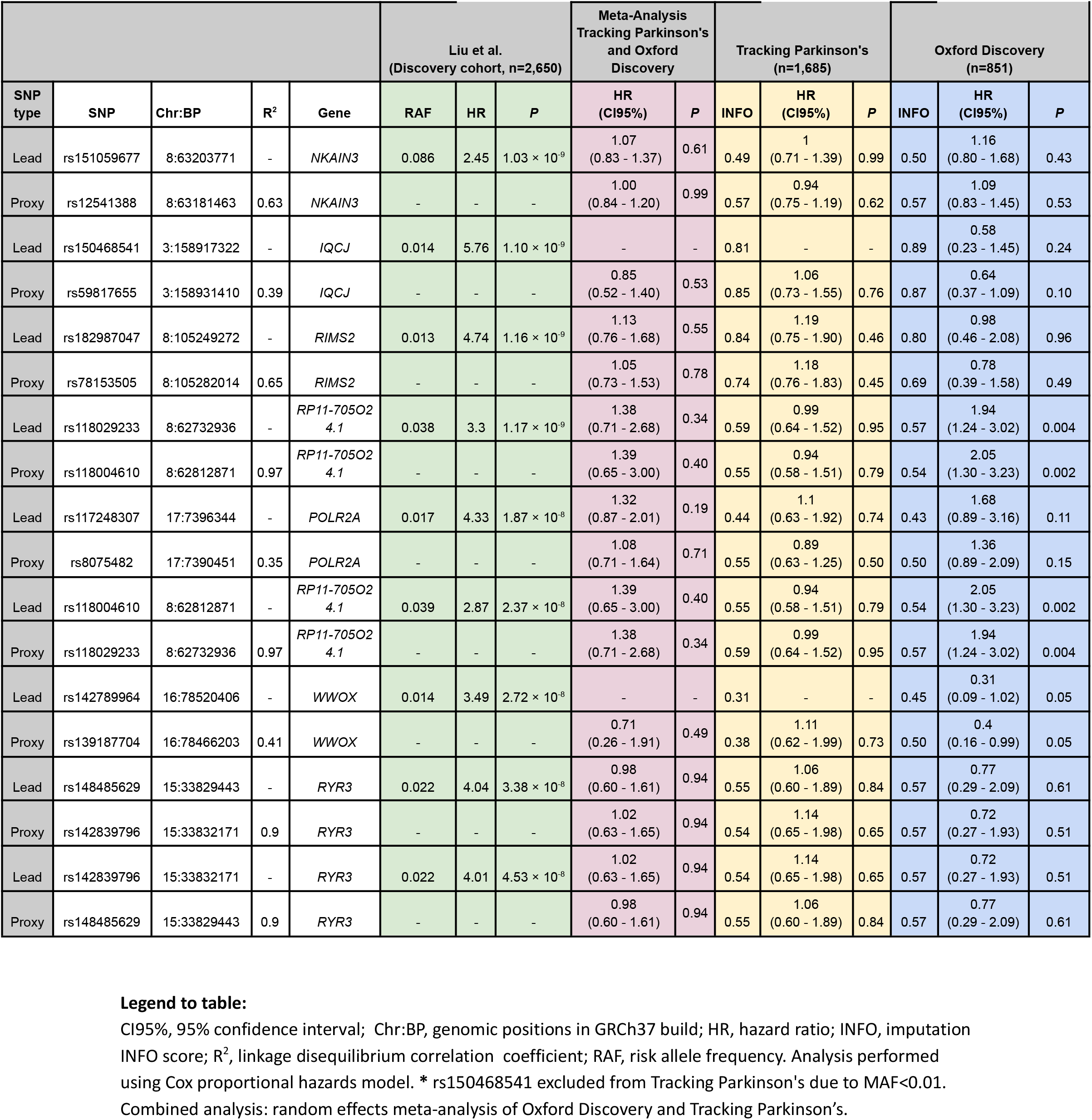
Survival analysis of nine candidate variants on the risk of progression from PD to PDD in UK cohorts

|  |  |  |  |  | Liu et al.<br>(Discovery cohort, n=2,650) |  |  | Meta-Analysis<br>Tracking Parkinson's<br>and Oxford<br>Discovery |  | Tracking Parkinson's<br>(n=1,685) |  |  | Oxford Discovery<br>(n=851) |  |  |
| --- | --- | --- | --- | --- | --- | --- | --- | --- | --- | --- | --- | --- | --- | --- | --- |
| SNP<br>type | SNP | Chr:BP | R<sup>2</sup> | Gene | RAF | HR | P | HR<br>(CI95%) | P | INFO | HR<br>(CI95%) | P | INFO | HR<br>(CI95%) | P |
| Lead | rs151059677 | 8:63203771 | - | NKAIN3 | 0.086 | 2.45 | 1.03 × 10<sup>-9</sup> | 1.07<br>(0.83 - 1.37) | 0.61 | 0.49 | 1<br>(0.71 - 1.39) | 0.99 | 0.50 | 1.16<br>(0.80 - 1.68) | 0.43 |
| Proxy | rs12541388 | 8:63181463 | 0.63 | NKAIN3 | - | - | - | 1.00<br>(0.84 - 1.20) | 0.99 | 0.57 | 0.94<br>(0.75 - 1.19) | 0.62 | 0.57 | 1.09<br>(0.83 - 1.45) | 0.53 |
| Lead | rs150468541 | 3:158917322 | - | IQCJ | 0.014 | 5.76 | 1.10 × 10<sup>-9</sup> | - | - | 0.81 | - | - | 0.89 | 0.58<br>(0.23 - 1.45) | 0.24 |
| Proxy | rs59817655 | 3:158931410 | 0.39 | IQCJ | - | - | - | 0.85<br>(0.52 - 1.40) | 0.53 | 0.85 | 1.06<br>(0.73 - 1.55) | 0.76 | 0.87 | 0.64<br>(0.37 - 1.09) | 0.10 |
| Lead | rs182987047 | 8:105249272 | - | RIMS2 | 0.013 | 4.74 | 1.16 × 10<sup>-9</sup> | 1.13<br>(0.76 - 1.68) | 0.55 | 0.84 | 1.19<br>(0.75 - 1.90) | 0.46 | 0.80 | 0.98<br>(0.46 - 2.08) | 0.96 |
| Proxy | rs78153505 | 8:105282014 | 0.65 | RIMS2 | - | - | - | 1.05<br>(0.73 - 1.53) | 0.78 | 0.74 | 1.18<br>(0.76 - 1.83) | 0.45 | 0.69 | 0.78<br>(0.39 - 1.58) | 0.49 |
| Lead | rs118029233 | 8:62732936 | - | RP11-705O2<br>4.1 | 0.038 | 3.3 | 1.17 × 10<sup>-9</sup> | 1.38<br>(0.71 - 2.68) | 0.34 | 0.59 | 0.99<br>(0.64 - 1.52) | 0.95 | 0.57 | 1.94<br>(1.24 - 3.02) | 0.004 |
| Proxy | rs118004610 | 8:62812871 | 0.97 | RP11-705O2<br>4.1 | - | - | - | 1.39<br>(0.65 - 3.00) | 0.40 | 0.55 | 0.94<br>(0.58 - 1.51) | 0.79 | 0.54 | 2.05<br>(1.30 - 3.23) | 0.002 |
| Lead | rs117248307 | 17:7396344 | - | POLR2A | 0.017 | 4.33 | 1.87 × 10<sup>-8</sup> | 1.32<br>(0.87 - 2.01) | 0.19 | 0.44 | 1.1<br>(0.63 - 1.92) | 0.74 | 0.43 | 1.68<br>(0.89 - 3.16) | 0.11 |
| Proxy | rs8075482 | 17:7390451 | 0.35 | POLR2A | - | - | - | 1.08<br>(0.71 - 1.64) | 0.71 | 0.55 | 0.89<br>(0.63 - 1.25) | 0.50 | 0.50 | 1.36<br>(0.89 - 2.09) | 0.15 |
| Lead | rs118004610 | 8:62812871 | - | RP11-705O2<br>4.1 | 0.039 | 2.87 | 2.37 × 10<sup>-8</sup> | 1.39<br>(0.65 - 3.00) | 0.40 | 0.55 | 0.94<br>(0.58 - 1.51) | 0.79 | 0.54 | 2.05<br>(1.30 - 3.23) | 0.002 |
| Proxy | rs118029233 | 8:62732936 | 0.97 | RP11-705O2<br>4.1 | - | - | - | 1.38<br>(0.71 - 2.68) | 0.34 | 0.59 | 0.99<br>(0.64 - 1.52) | 0.95 | 0.57 | 1.94<br>(1.24 - 3.02) | 0.004 |
| Lead | rs142789964 | 16:78520406 | - | WWOX | 0.014 | 3.49 | 2.72 × 10<sup>-8</sup> | - | - | 0.31 | - | - | 0.45 | 0.31<br>(0.09 - 1.02) | 0.05 |
| Proxy | rs139187704 | 16:78466203 | 0.41 | WWOX | - | - | - | 0.71<br>(0.26 - 1.91) | 0.49 | 0.38 | 1.11<br>(0.62 - 1.99) | 0.73 | 0.50 | 0.4<br>(0.16 - 0.99) | 0.05 |
| Lead | rs148485629 | 15:33829443 | - | RYR3 | 0.022 | 4.04 | 3.38 × 10<sup>-8</sup> | 0.98<br>(0.60 - 1.61) | 0.94 | 0.55 | 1.06<br>(0.60 - 1.89) | 0.84 | 0.57 | 0.77<br>(0.29 - 2.09) | 0.61 |
| Proxy | rs142839796 | 15:33832171 | 0.9 | RYR3 | - | - | - | 1.02<br>(0.63 - 1.65) | 0.94 | 0.54 | 1.14<br>(0.65 - 1.98) | 0.65 | 0.57 | 0.72<br>(0.27 - 1.93) | 0.51 |
| Lead | rs142839796 | 15:33832171 | - | RYR3 | 0.022 | 4.01 | 4.53 × 10<sup>-8</sup> | 1.02<br>(0.63 - 1.65) | 0.94 | 0.54 | 1.14<br>(0.65 - 1.98) | 0.65 | 0.57 | 0.72<br>(0.27 - 1.93) | 0.51 |
| Proxy | rs148485629 | 15:33829443 | 0.9 | RYR3 | - | - | - | 0.98<br>(0.60 - 1.61) | 0.94 | 0.55 | 1.06<br>(0.60 - 1.89) | 0.84 | 0.57 | 0.77<br>(0.29 - 2.09) | 0.61 |

There are a number of possible reasons which may explain why we have failed to detect an effect of the *RIMS2* locus on PD dementia in our study, using a very similar methodology to Liu and colleagues study in two large well-powered UK cohorts.

Genotyping and imputation accuracy may be important, particularly when dealing with rare alleles. In our replication study the *RIMS2* variant and its’ proxy are imputed to a high quality. Although the study of Liu and colleagues is large it has limited power to detect an effect of rarer alleles on dementia. If 30% of their 4,600 cases develop dementia then 80% power will be reached with an effect estimate (beta) of 0.6, at a similar effect allele frequency under an additive model. If their finding is correct then it is likely to be subject to a major “winner’s curse” effect, that is that the case/control or allele related survival differences have been identified at the upper limits of the underlying population distribution and so large follow-on studies will be needed to replicate these findings and to verify the true, likely smaller effects.

It is possible, particularly in analyses of rare alleles, that there may be population specific effects so that a true effect may be detectable only in specific studies / populations, related to variation in background allele frequency. In addition, confounding by population stratification may occur particularly as the frequency of rare variants in *GBA*, associated with dementia, vary between Northern European populations and are most common in Ashkenazi Jewish PD patients. However, the *RIMS2* risk allele appears to have a similar allele frequency in Finnish, Ashkenazi Jewish and non-Finnish European populations (GnomAD) so currently there is no support for major population stratification confounding effects.

The individual study statistics are not reported by Liu and colleagues, so study heterogeneity is difficult to evaluate, but their paper involves 15 different cohorts from Europe and North America. Given a risk allele frequency of 1.3% and the individual cohort sizes, the number of risk allele carriers per study can be predicted to lie between 2 and 8 suggesting that there are likely to be wide individual study confidence intervals in some studies. It would be interesting to know if the effect is driven by specific cohorts and to assess their relative contribution to the *RIMS2* discovery. Potentially, varying definitions of dementia superimposed on underlying variation in the *RIMS2* risk allele frequency between cohorts may confound their analysis. Liu and colleagues report both a lambda and linkage disequilibrium score intercept of >1.05 implying that there is potential inflation of summary statistics and underlying study heterogeneity. We would recommend the use of genomic control to help reduce the impact of these factors both on the cohort and meta-analytic level. Given the large number of small contributing cohorts, we would have liked to have seen a forest plot showing the effect estimates for each individual cohort as well as their weights and measures of heterogeneity (I^2^) to demonstrate the relative contribution of the individual studies to their pooled analysis.

Finally, misdiagnosis is a potential source of error and the Tracking and Oxford discovery cohorts have been screened longitudinally for the emergence of atypical parkinsonism and alternative diagnoses at serial study visits. However, *RIMS2* is not a known risk locus for DLB, PSP, CBD or MSA (albeit in common variant studies for PSP, CBD and MSA) so confounding by misdiagnosis seems less likely (6–9).

In conclusion, the definition of modifying factors in PD progression is likely to be vitally important in understanding disease biology and developing new disease modifying treatments. Liu and colleagues have developed and analysed an important large collaborative dataset which advances the field, in conjunction with the previous work on PD progression. However, we are not able to replicate the *RIMS2* locus in large well powered UK PD progression studies and currently *RIMS2* should be considered as a possible rather than a definite risk locus for PD dementia.

## Data Availability

Data availability:
The original data used in this study is available from the Tracking Parkinson’s (www.trackingparkinsons.org.uk) and Oxford Discovery teams (www.dpag.ox.ac.uk/opdc)
Code and method availability:
The analysis protocol, code and full summary statistics are available at GitHub (https://github.com/huw-morris-lab/dementia_GWAS_replication;
doi: 10.5281/zenodo.5137374)

https://doi.org/10.5281/zenodo.5137374

## Cohort studies

Both Tracking Parkinson’s and Oxford Discovery are primarily funded and supported by Parkinson’s UK. Both studies are supported by the National Institute for Health Research (NIHR) Dementias and Neurodegenerative Diseases Research Network (DeNDRoN). Oxford Discovery is also supported by the NIHR Oxford Biomedical Research Centre based at Oxford University Hospitals NHS Trust, and the University of Oxford. This research was supported by the National Institute for Health Research University College London Hospitals Biomedical Research Centre. The UCL Movement Disorders Centre is supported by the Edmond J. Safra Philanthropic Foundation.

## Genetic analysis

Work on the genetics of progression in Parkinson’s is supported by Parkinson’s UK (PhD Studentship to Dr Tan H-1703, Understanding and predicting Parkinson’s progression) and by Aligning Science Across Parkinson’s [Grant ID: ASAP 0478] through Michael J. Fox Foundation for Parkinson’s Research (MJFF) (Aligning Science Across Parkinson’s (ASAP) Collaborative Research Network, Chevy Chase, MD, 20815). For the purpose of open access, the author has applied a CC BY public copyright license to all Author Accepted Manuscripts arising from this submission.

## Competing interests

M.M.X.T is supported by Parkinson’s UK

M.A.N.’s participation in this project was part of a competitive contract awarded to Data Tecnica International LLC by the National Institutes of Health to support open science research, he also currently serves on the scientific advisory board for Clover Therapeutics and is an advisor to Neuron23 Inc as a data science fellow. This research was supported in part by the Intramural Research Program of the NIH, National Institute on Aging (NIA), National Institutes of Health, Department of Health and Human Services; project number ZO1 AG000535, as well as the National Institute of Neurological Disorders and Stroke.

J.H. is supported by the UK Dementia Research Institute, which receives its funding from DRI Ltd, funded by the UK Medical Research Council, Alzheimer’s Society, and Alzheimer’s Research UK. He is also supported by the MRC, Wellcome Trust, Dolby Family Fund, National Institute for Health Research University College London Hospitals Biomedical Research Centre.

Y.B.-S. has received grant funding from the MRC, NIHR, Parkinson’s UK, NIH, and ESRC.

N.M.W. is supported by Parkinson’s UK.

M.T.H. receives grants from Parkinson’s UK, Oxford NIHR Biomedical Research Centre, and MJFF and is an adviser to the Roche Prodromal Advisory and Biogen Digital advisory boards.

D.G.G. has received grants from Michael’s Movers, the Neurosciences Foundation, and Parkinson’ s UK, and honoraria from BIAL Pharma, GE Healthcare, and consultancy fees from Acorda Therapeutics and the Glasgow Memory Clinic..

H.R.M. reports paid consultancy from Roche. Research Grants from Parkinson’s UK, Cure Parkinson’s Trust, PSP Association, CBD Solutions, Drake Foundation, Medical Research Council, Michael J Fox Foundation. Dr Morris is a co-applicant on a patent application related to C9ORF72 - Method for diagnosing a neurodegenerative disease (PCT/GB2012/052140)

All other authors did not declare any funding sources that directly contributed to this study.

M.M.X.T. takes responsibility for the integrity of the data and the accuracy of the data analysis.

## Author contribution

HRM and MMXT conceived, obtained funding for and designed the genetic study.

DGG, MTH, Y B-S, TF, NWW, NMW, JH and HRM conceived and led the longitudinal clinical cohort and biomarker studies.

JH, MS, RR, MAL, Y B-S, NMW, MTH, MAN, DG contributed to the study design and interpretation MAL and Y B-S led the clinical cohort data management and analysis.

MMXT, RR and MS carried out the statistical and genetic analyses. CB, LH carried out the genotyping led by NMW in his laboratory.

Patient samples and phenotypic data were collected by MMXT, TF, MTH, DGG and HRM HRM, MMXT, RR and MS drafted the manuscript.

All authors reviewed, edited and approved the manuscript prior to submission.

## Data availability

The original data used in this study is available from the Tracking Parkinson’s (www.trackingparkinsons.org.uk) and Oxford Discovery teams (www.dpag.ox.ac.uk/opdc)

## Code and method availability

The analysis protocol, code and full summary statistics are available at GitHub (https://github.com/huw-morris-lab/dementia_GWAS_replication; doi: 10.5281/zenodo.5137374)

## Clinical studies

Tracking Parkinson’s has multi-centre research ethics approval West of ScotlandResearch Ethics Committee: IRAS 70980, MREC 11/AL/0163, Clinicaltrials.gov: NCT02881099). Oxford Discovery has multi-centre research ethics approval (South Central Oxford A Research Ethics Committee 16/SC/0108). Each subject provided written informed consent for participation.

## References

1. Liu, G. et al. Genome-wide survival study identifies a novel synaptic locus and polygenic score for cognitive progression in Parkinson’s disease. Nat. Genet. 1–7 (2021).

2. Tan, M. M. X. et al. Genome-Wide Association Studies of Cognitive and Motor Progression in Parkinson’s Disease. Mov. Disord. (2020). doi:10.1002/mds.28342

3. Iwaki, H. et al. Genomewide association study of Parkinson’s disease clinical biomarkers in 12 longitudinal patients’ cohorts. Mov. Disord. 34, 1839–1850 (2019).

4. Malek, N. et al. Tracking Parkinson’s: Study Design and Baseline Patient Data. J. Parkinsons. Dis.5, 947–959 (2015).

5. Szewczyk-Krolikowski, K. et al. The influence of age and gender on motor and non-motor features of early Parkinson’s disease: initial findings from the Oxford Parkinson Disease Center (OPDC) discovery cohort. Parkinsonism Relat. Disord. 20, 99–105 (2014).

6. Sailer, A. et al. A genome-wide association study in multiple system atrophy. Neurology 87, 1591–1598 (2016).

7. Kouri, N. et al. Genome-wide association study of corticobasal degeneration identifies risk variants shared with progressive supranuclear palsy. Nat. Commun. 6, 7247 (2015).

8. Höglinger, G. U. et al. Identification of common variants influencing risk of the tauopathy progressive supranuclear palsy. Nat. Genet. 43, 699–705 (2011).

9. Chia, R. et al. Genome sequencing analysis identifies new loci associated with Lewy body dementia and provides insights into its genetic architecture. Nat. Genet. 53, 294–303 (2021).

